# How Many Hours Do Internal Medicine Residents At University Of Toronto Spend Onboarding At Hospitals Each Year? A Cross-sectional Survey Study

**DOI:** 10.1101/2022.06.10.22276103

**Authors:** Benazir Hodzic-Santor, Varuna Prakash, Ashley Raudanskis, Edmund Lorens, Allan S. Detsky, Michael Fralick

## Abstract

**Background:** Burnout among medical residents is common. One source of burnout is the need to complete redundant administrative tasks such as onboarding processes at various hospitals.

**Objectives:** To quantify the time residents at the University of Toronto spend onboarding at teaching hospitals, to identify areas of redundancy in onboarding processes, and to identify trainee perceptions of onboarding processes.

**Methods:** We conducted a quality improvement survey of core internal medicine residents at the University of Toronto where residents rotate through multiple different teaching hospitals. The primary outcome was time spent onboarding. Secondary outcomes included perceptions of the onboarding process, and impact on well-being.

**Results:** 41% (N=93) of all Internal Medicine residents completed the survey. Most (n=81, 87%) rotated through at least four hospitals and 24 (26%) rotated through more than 5 in the preceding year. The median number of hours spent on the onboarding process was 5 hours per hospital (IQR 1-8) and these tasks were often completed when trainees were post-call (82%, n=76) or outside of work hours (97%, n= 90). The cumulative number of hours spent each year on onboarding tasks by the 93 trainees was 2325 hours (97 days) which extrapolates to 5625 hours (234 days) for all 225 trainees in the core internal medicine program. Most residents reported high levels of redundancy across hospital sites (n=79, 85%) and felt that their well-being was negatively affected (73%, n=68).

**Conclusions:** The median internal medicine resident at the University of Toronto spent 5 hours onboarding for each hospital. There is considerable redundancy and the process contributes to self-reported burnout.

## INTRODUCTION

Burnout among resident physicians is a well-recognized phenomenon that has received increasing attention in recent years. The burnout syndrome is now included as an occupational condition in the World Health Organization’s *International Disease Classification (ICD) 11*, and comprises feelings of energy depletion or exhaustion, negativity or cynicism towards one’s job, and reduced professional efficacy^1^.The literature suggests that burnout rates range from 15% to 75%^2,3^. Root causes of burnout can be broadly categorized into personal and demographic factors (e.g., gender, family structure), programmatic structure of the training experience (e.g., work hours, frequency of on-call), work-related stress, and interpersonal workplace relationships, to name a few^2–6^. Interventions to reduce burnout span a spectrum of individual-based solutions (e.g., wellness sessions, exercise programs, mindfulness) and workplace solutions (e.g., work hour restrictions, mentorship fatigue management, workload balancing)^3^. The effectiveness of these interventions on trainee burnout is poorly studied.

Given the known impact of physician burnout on rates of depression and suicidal ideation, physical health status, and job satisfaction, there is a strong need to identify and mitigate factors contributing to burnout. One aspect of physicians’ work that has received relatively less attention is the burden of non-clinical and non-patient facing administrative tasks. For example, there is growing recognition that documentation and administrative requirements associated with Electronic Medical Records (EMRs) contribute significantly to physician burnout^7–9^. A recent US-based national survey found that physician task load – including documentation, computerized order entry, and completion of mandatory e-learning modules – was associated with an increase in burnout^10,11^, and suggested that as much as a 10% drop in task load could reduce odds of burnout by a third^11^.

Training programs that include more than one hospital require that residents learn multiple EMRs, and navigate multiple and often redundant onboarding processes. The Internal Medicine training program at the University of Toronto includes training at five core affiliated hospitals and is Canada’s largest program. The objective of this study was to quantify the amount time internal medicine trainees spend onboarding at our affiliated hospitals, to identify areas of redundancy in onboarding processes, and to identify trainee perceptions of onboarding processes.

## METHODS

### Study Setting, Data Source, Study Population

We surveyed internal medicine residents at the University of Toronto in Toronto, Ontario in post-graduate years (PGY) 1, 2 and 3 through a web-based survey disseminated via email and social media in May-June 2021. Study participation was voluntary, and respondents were provided with a small coffee card as compensation for their time. Upon review of the project, the Mount Sinai Hospital Research Ethics Board determined that this activity is a Quality Improvement initiative and does not constitute research under Chapter 2 of the Tri-Council Policy Statement 2: Ethical Conduct for Research Involving Humans (TCPS2), and therefore does not fall within the scope of REB review.

The Internal Medicine residency at the University of Toronto is a single program with central recruitment and administration. There are 5 major teaching hospitals and 3 core affiliated community hospitals that offer training to the residents over 3 core years (PGY1, PGY2 and PGY3). As part of their clinical training, residents rotate through several hospitals (up to 6 in the same academic year), usually for a month at a time, but sometimes for shorter periods. Because each hospital is an individual institution with separate governance and administration, they each have different EMR technology and unique administrative and onboarding requirements that residents must complete before they are granted privileges to begin their rotation.

The onboarding processes typically include e-learning modules covering health and safety, privacy, professional behavior, and security, as well as steps to acquire identification badges, hospital scrubs and pagers. As a result of this structure (a single program and many independent teaching hospitals) residents must complete the onboarding process several times each year. The certification only lasts 1 year, and so must be repeated for the second and third years of training.

### Survey Creation and Content

Our study team drafted an initial set of survey questions that was beta-tested by chief medical residents at our hospitals. Iterative changes to survey structure and wording were made based on their feedback. The final survey included a free-text comment box and 10 questions that captured the following areas: demographics, number of hospitals where the resident provided clinical care in the preceding year, onboarding process at each hospital, time spent on the onboarding process, experience with the onboarding process, degree of redundancy across hospitals, perceived value of the onboarding process, and impact of the process on their well-being. We considered the following tasks to be part of the onboarding process: electronic medical record training, occupational health and safety modules, privacy and other professional behavior modules, acquiring a hospital badge, acquiring a hospital pager, acquiring scrubs, and miscellaneous modules (e.g. computer safety).

### Outcomes

Our primary outcome was the time (in hours) internal medicine trainees spend onboarding at teaching hospitals per year. We also identified areas of redundancy, and characterized trainee perceptions of onboarding processes.

### Statistical analysis and language processing

Descriptive statistics were used to summarize the quantitative data. Natural language processing and manual review by our study team was used to summarize the free-text data; specifically, a combination of regular expression and sentiment analysis was performed. Sentiment analysis was conducted using three different transformer-based models, a neural network-based technique for natural language processing. The models employed have been pre-trained on a large corpus of English text in a self-supervised fashion and have routinely been applied in sentiment analysis^12^. Models 1 and 2 used BERT, a bidirectional transformer model pretrained using masked language modeling and next sentence prediction^13^. Model 3 used RoBERTa, a transformer-based model that builds on BERT by removing the next-sentence pretraining objective, and training with much larger mini-batches and learning rates^14^.

Model 1 classifies responses into either positive or negative whereas model 2 predicts the sentiment of the response as the number of stars (between 1 and 5). Model 3 classified responses as being either positive (2), neutral (1), or negative (0). To help contextualize the results from natural language processing of the free text comments from our survey, we ran our models on a separate dataset of teacher effectiveness scores completed by internal medicine residents from 2020-2021. These data included anonymized free text comments from 138 residents, including the same cohort that completed our survey.

The most common sentiments were identified using the Bing Liu sentiment lexicon of the tidytext package. This lexicon contains about 2006 positive words and 4683 negative words and includes misspellings, slang words and morphological variants. For all positive and negative words identified in the free-text responses, we extracted the surrounding 4-6 words to investigate if the words before or after altered the meaning. All analyses were performed using R Statistical Software (version 4.0.5; R Foundation for Statistical Computing, Vienna, Austria).

## RESULTS

### Respondent Demographics

Of the 225 residents who were emailed the survey, 93 completed the survey (41%). Respondents evenly represented all years of training, with approximately 34% (n=32) in PGY1, 32% (n=30) in PGY2, and 30% (n=28) in PGY3. The majority (n=81; 87%) of trainees rotated through at least four hospitals and 24 (26%) rotated through more than 5 in the preceding year.

### Onboarding Processes

Most residents were asked to complete the same components of the onboarding process at 3 or more hospitals. This included completing online modules, EMR training, and obtaining a pager, badge, and scrubs (Figure 1). The median number of hours spent on the onboarding process at each hospital was 5 (IQR 1-8) and most often these tasks were completed when trainees were post-call (82%, n=76) or outside of work hours (e.g., vacation, evenings, weekends; 97%, n= 90).

**Figure 1:**
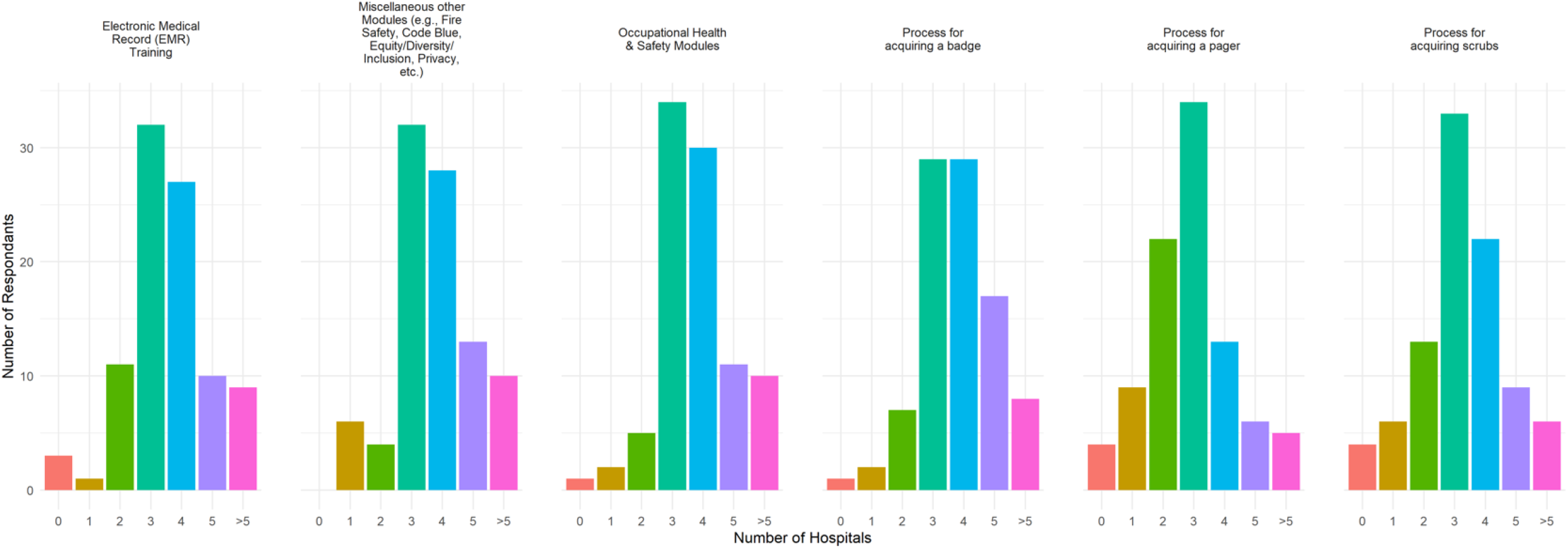
Resident responses on number of hospitals requiring the completion of various onboarding components Legend: miscellaneous modules include fire safety, privacy, equity diversity and inclusion, etc. EMR = electronic medical record.

To estimate the total amount of time all trainees who replied to our survey spent on the onboarding process, we multiplied the median number of reported hours by the median number of hospitals rotated through each year by the number of trainees who completed the survey. Based on this, the total number of hours spent by all trainees each year on onboarding tasks was 2325 hours (97 days or 14 weeks) with the average resident spending 25 hours per year completing onboarding processes. The cumulative number of hours spent each year on onboarding tasks extrapolates to 5625 hours (234 days or 33 weeks) for all 225 trainees in the core internal medicine program.

Redundancy was assessed by asking participants to estimate the percentage of content that was duplicated or redundant across sites (0-20%, 21-40%, 41-60%, 61-80% or 81-100%). 85% of respondents (n=79) felt that >60% of content was redundant, and 97% (n=90) felt >40% was redundant (Table 1).

**Table 1:** Estimated Percentage of onboarding module content that was duplicated or redundant across sites

| Percentage of duplicated and/or redundant content | Number of respondents |
| --- | --- |
| 0 - 20% | 1 |
| 21 - 40% | 2 |
| 41 - 60% | 11 |
| 61 - 80% | 39 |
| 81 - 100% | 40* |

\* i.e., 40 residents felt that 81-100% of the module content was duplicated or redundant across sites.

### Attitudes towards Onboarding Processes

Residents were also asked to select the adjectives that best described their feelings towards the onboarding process (Figure 2). The most frequently endorsed adjective was “annoyed” (77%, n=72). Most residents also described the process as boring, exhausting, frustrating, and unproductive. Fewer than 10% described that the process made them feel informed, competent, or prepared.

**Figure 2:**
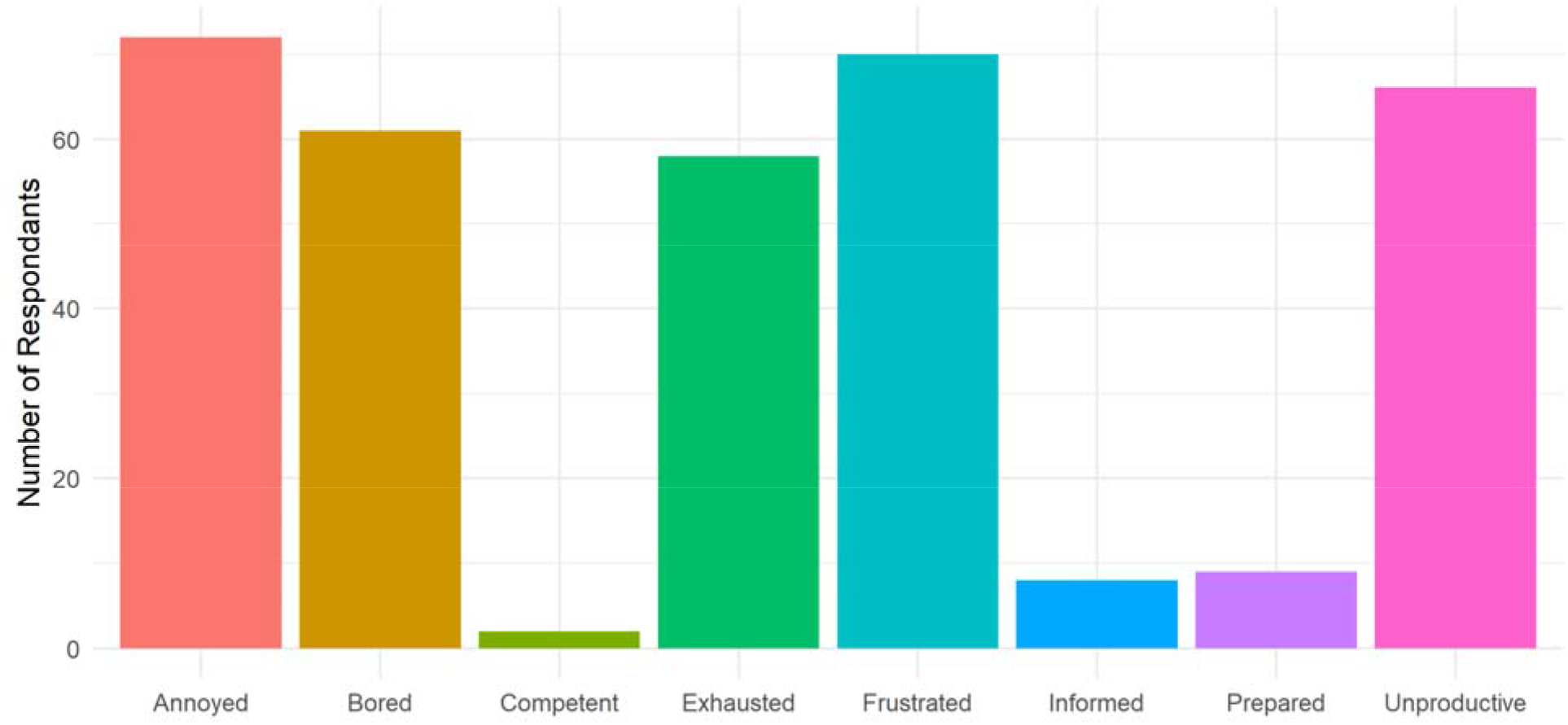
Adjectives chosen by residents to describe the onboarding process

The majority of residents described that the onboarding process worsened their well-being (73%, n=68). Residents at higher levels of training more frequently stated that their well-being was negatively affected; 63% (n=20) of PGY1 residents, 75% (n=24) of PGY2 residents and 86% (n=24) of PGY3 residents agreed that their well-being was worsened. Overall, the impact of the onboarding process was described as irritating by most residents; 49% (n=46) described it as a major irritant and 46% (n=43) described it as a minor irritant. Only 2% (n=2) felt the process was a beneficial way to learn important material, or an essential process that guaranteed their success on the job. When asked specifically about the online onboarding modules (72%, n= 67) felt completing modules was not a good use of time. Most residents reported that they completed the end of module quizzes without reading the content (69%, n=64). Only 22% (n=20) of respondents felt they could remember the module content 6 months later, with residents in PGY1 reporting increased retention (31%, n=10) compared to those in PGY2 (20%, n=6) or PGY3 (14%, n=4).

Of the respondents, 35% (n=33) provided additional comments in the free-text box. Only comments pertaining to onboarding were included in the sentiment analysis (n=32), with one comment thanking the team for creating the survey excluded from further analysis. Sentiment analysis found that 88% of the comments expressed predominantly negative emotions (Table 2). When rated on a scale of 1 to 5, with 1 being the most negative and 5 being the most positive, 61% (n =20) of the comments were rated as either 1 or 2 stars. The most common sentiment words and expressions were “redundant/redundancy” (n=11), “no protected time/lack of protected time/should have protected time” (n=6), “frustrate/frustrated/frustrating” (n=4) and “annoy/annoyed/annoying” (n=4). Although words like “effective” and “helpful” appeared, they were not used to describe the current onboarding process in a positive sense. For example: “it would be best to choose which module is more *effective*” or “EMR training is *helpful* except when we have done it before”. The frequencies of common sentiment words are summarized in Figure 3.

**Table 2:** Sentiment analysis of onboarding survey and teacher effectiveness comments

|  | **Module comments**<br>**(N=33)** | **Teacher comments**<br>**(N=138)** |
| --- | --- | --- |
| Model 1: 5 star: 4 star : 3 star : 2 star : 1 star [model confidence] | 2:3:8:13:7 [48%] | 95:33:5:5 [69%] |
| Mode 2: % of comment that are negative [model confidence] | 12% [99%] | 89% [99%] |
| Model 3: mean score [model confidence confidence] | 0.5 [75%] | 1.86 [90%] |

Model 1 characterizes the free text comments on a scale of 1 star (highly negative) to 5 stars (highly positive). Model 2 characterized the percentage of comments that were negative using a previously validated lexicon. Model 3 calculates a mean score of free text comments ranging from 0 (negative) to 2 (positive).

**Figure 3:**
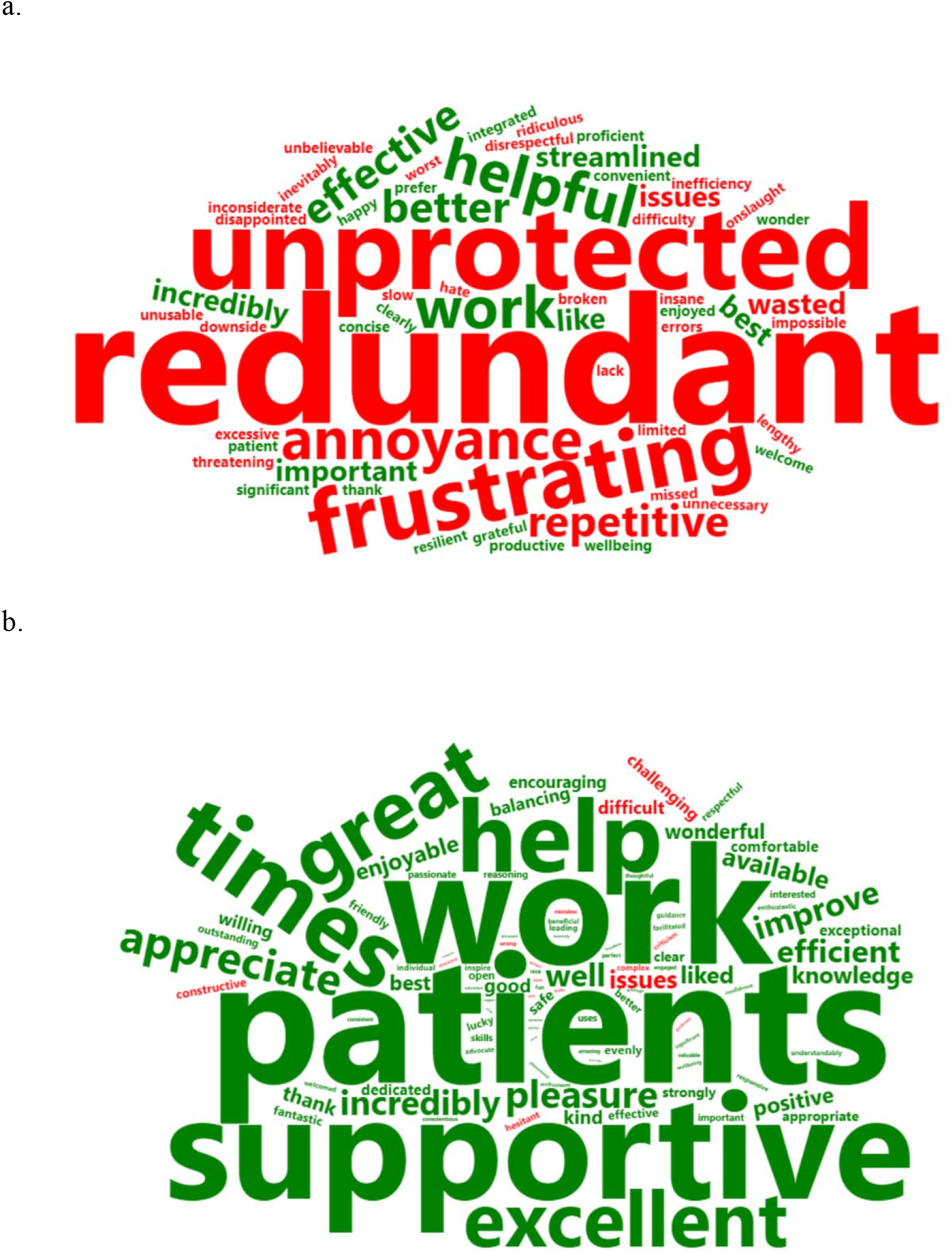
Summary of words used by residents to describe the onboarding process (a) and words used in teacher evaluations (b)

To provide a comparison for context, we preformed the same analysis on internal medicine attending staff teacher effectiveness feedback comments from a random sample of comments from 139 residents after each rotation during the same time period that our survey was administered. One comment was excluded due to its excessive length. Sentiment analysis found that 89% of the comments reflected predominantly positive emotions (Table 2). In contrast to the results of our onboarding survey, only 7% (n =10) of teacher reviews were rated as 1 or 2 stars.

## DISCUSSION

This quality improvement study involving a survey of internal medicine residents at our academic program identifies that current onboarding processes impose a significant time burden on trainees, suffer from content redundancy, and contribute to trainee frustration and possible burnout.

Due to the number of independent training sites at our program, most of our respondents were required to complete onboarding processes at four or more sites during the academic year, with most sites containing the same core elements of onboarding processes. Based on self-reported data, the 93 trainees who responded to the survey cumulatively spent a staggering 2325 hours (i.e., the equivalent of 3.2 months) on various elements of administrative onboarding tasks; extrapolating these results to our entire group of residents produces an estimate of 8.2 months.

Put another way, this cumulative time spent on onboarding tasks by all trainees in our program is the equivalent of 2/3 of a year of academic residency training for one person. A vast majority of trainees completed these onboarding tasks during non-working times (e.g., post-call, on weekends, or on vacation); it is therefore unsurprising that most trainees we surveyed indicated that these processes were a ‘major irritant’ and had a negative impact on their well-being. These findings suggest that decreasing the time burden of onboarding processes and providing reasonable protected time to complete these processes may have a direct positive impact on trainees’ workload and well-being.

Our results also highlight the redundancy in onboarding processes across multiple institutions, which is a result of separate administrations that each have their own credentialing processes (and EMR systems). While standardizing EMR systems across all these hospitals is unlikely to happen in the near future due to institutional factors, other onboarding processes (e.g., badge access, completion of mandatory modules) are less institution-specific and could in theory be centralized more easily if there is sufficient motivation to make this change.

Furthermore, responses to our survey also indicated that the e-learning modules may be ineffective methods of imparting information. Most trainees reported ‘clicking through’ modules without absorbing the content, with poor recall of content 6 months later. These results suggest that centralizing mandatory modules across hospitals, especially for content that is not institution specific (e.g., privacy modules), may significantly improve trainees’ experience. Additionally, these results suggest a strong need to re-examine the content, user engagement, and didactic efficacy of these modules to ensure that they are achieving their intended goal.

We conducted a sentiment analysis using three different advanced natural language processing models and also ran these models on a separate dataset of teacher evaluation comments to help contextualize the sentiment of the comments from our survey. The results were incredibly negative and this was consistently observed across all three models used. These results are congruent with literature suggesting that increasing burden of physician tasks, especially those that are clerical and administrative in nature, are directly associated with clinician burnout^10^. We hope to convey a ‘burning platform’ for change with the results of our study, and suggest that there is considerable potential to directly improve the trainee experience and minimize contributors to burnout by overhauling existing onboarding processes and making them less burdensome.

In calling for change, we recognize that overhauling deeply entrenched processes is challenging, particularly at large and distributed institutions such as ours, where the people in charge of credentialing physicians are more concerned about risk management than physician well-being. In our appendix we provide potential solutions and categorize them as “low effort solutions” that could be implemented with minimal work and “high effort solutions” that require a significant amount of time, effort, money and coordination.

### Limitations

Our study has limitations. Firstly, this study focused on a single specialty of residency in one university program with independent training sites that currently have very different onboarding processes. As such, these results may not reflect the experiences of trainees at other institutions or training programs. Future work could expand on this proof-of-concept by broadening the sample population across institutions and specialties. Secondly, our methodology relied on self-reported data. For instance, trainees were asked to self-estimate the amount of time they spent on onboarding, because the actual onboarding completion time is not recorded. Similarly, redundancy in onboarding modules was directly recalled by trainees and not objectively verified. While this self-reporting provides valuable insights into trainees’ perceptions of the burden of onboarding, we acknowledge that it is subject to estimation bias. In future, a comparison of subjective and objective estimates of metrics such as hours spent on onboarding and percentage of content redundancy could yield interesting insights into the effectiveness of onboarding modules and processes. Finally, while our response rate of 41% is consistent with other published studies that surveyed trainees, this may have resulted in selection bias and thus potentially an over-estimation of the time spent on the onboarding tasks.

## Conclusion

Burnout is common in residency, and may be worsened by the current approach of hospital on-boarding. Our results show that onboarding processes are an overlooked component of the trainee experience, and lead to significant time burden, negative impact on trainee well-being, and increase in trainee burnout. Redundant and excessively frequent onboarding processes are a prime example of costs without benefit. Interventions such as harmonizing onboarding processes across sites, providing protected time to complete onboarding, and increasing the efficacy of onboarding content are necessary to reduce trainee burnout and improve well-being.

## Data Availability

All data produced in the present work are contained in the manuscript.

### Appendix

**Appendix Table 1.** Low effort solutions to improving the onboarding process

|  | **Suggested approach** | **Current approach** |
| --- | --- | --- |
| 1 | Allow hospital ID badge to be active for 3-years to cover their 3 years of GIM. | Most hospitals issue a badge for a 1-year time period, necessitating an updated badge each year. |
| 2 | Allow access to an electronic medical record for 3-years to cover their 3 years of GIM. | Most hospitals only allow 1-year of access at a time necessitating trainees to repeat the process of getting access to the electronic medical record |
| 3 | Hospitals accept a certificate from PGME showing completion of standard onboarding modules (e.g., WHIMIS, hand hygiene). | Trainees complete modules through PGME and each hospital they rotate through. |
| 4 | Provide protected time for trainees to complete the onboarding during an academic half-day in July and a second half-day in January | 90% of trainees complete the onboarding process during post-call days or on evenings and weekends. |

**Appendix Table 2.** Higher effort solutions to overhaul the onboarding process at Toronto teaching hospitals

|  | **Suggested approach** | **Current approach** |
| --- | --- | --- |
| 1 | Provide a single badge that is recognized and accepted by all teaching hospitals. | At each hospital, trainees are required to get a unique ID badge. |
| 2 | Implement a single onboarding process, similar to what is available to staff physicians who work at multiple hospitals in Toronto. | Each hospital has their own onboarding process and there is no communication between sites about what trainees have done at other sites. |
| 3 | Have a single electronic medical record system at all Toronto hospitals. | Every hospital in Toronto has a different and unique electronic medical record system. |
| 4 | Remove the need for pagers; instead, residents can be paged to their cellphone, similar to what occurs with staff physicians. | Each individual hospital issues a unique pager. |

#### Survey

1. What is your current level of training?
  - Options: PGY1, PGY2, PGY3
2. How many different hospitals did you rotate through in the last 12 months? Note: Hospitals within the same health system are to be counted as separate hospitals (e.g. TGH and TWH are counted as two hospitals)
  - Options: 0, 1, 2, 3, 4, 5, >5
3. How many of the hospitals you indicated above required you to complete the following: Occupational Health & Safety Modules, Miscellaneous other Modules (e.g., Fire Safety, Code Blue, Equity/Diversity/Inclusion, Privacy, etc.), Electronic Medical Record (EMR) Training, Process for acquiring a badge, Process for acquiring a pager, Process for acquiring scrubs
  - Options provided for each item of onboarding: 0, 1, 2, 3, 4, 5, >5
4. On average, how many hours per hospital do you estimate it took to complete all applicable components of the above onboarding process? Please enter a number of hours.
  - Open-Ended Response
5. On average, which of the following statements apply to your completion of these onboarding components? Select all that apply.
  - I completed these components on my post-call days.
  - I completed these components outside of working hours (e.g., evenings, weekends) or while on vacation.
  - I had protected time while at work to complete these components.
6. What percent of the onboarding module content you completed do you feel was duplicated, redundant, or very similar across sites?
  - 0 - 20% was duplicated and/or redundant
  - 21 - 40% was duplicated and/or redundant
  - 41 - 60% was duplicated and/or redundant
  - 61 - 80% was duplicated and/or redundant
  - 81 - 100% was duplicated and/or redundant
7. Which of the following adjectives applies to how you felt about the onboarding processes you have had to complete? Select all that apply.
  - Annoyed
  - Bored
  - Exhausted
  - Frustrated
  - Unproductive
  - Informed
  - Competent
  - Prepared
  - Excited
  - Productive
  - Indifferent
  - Angry
  - Exasperated
8. Once you completed the modules (e.g., Handwashing, Code Blues, Fire Safety, Privacy), which of the following statements best describes how you felt? Select all that apply.
  - I learned something new through these modules
  - I felt more prepared for my clinical experiences
  - I felt I could remember the content of these modules 6 months later
  - I completed the module quizzes without reading the module content
  - I felt that completing the modules was not an effective way for me to learn the content
9. Which of the following statements best describes the impact of these onboarding processes on your well-being? The onboarding processes:
  - Worsened my well-being
  - Did not affect my well-being
  - Enhanced my well-being
10. Which of the following best describes the overall impact of these onboarding processes on your education? The onboarding process is:
  - A minor irritant
  - A major irritant
  - An essential process that guarantees my success on the job
11. Optional: Please provide any additional comments on your experience with the onboarding process/onboarding modules
  - Open-Ended Response

## Notes

### Competing Interest Statement

The authors have declared no competing interest.

### Funding Statement

This study did not receive funding.

### Author Declarations

Upon review of the project, the Mount Sinai Hospital Research Ethics Board has determined that this activity is a Quality Improvement initiative and does not constitute research under Chapter 2 of the Tri-Council Policy Statement 2: Ethical Conduct for Research Involving Humans (TCPS2), and therefore does not fall within the scope of REB review.

